# Dissecting the Genetic Architecture of Intracranial Aneurysms

**DOI:** 10.1101/2023.07.30.23293390

**Authors:** Shaunak S. Adkar, Julie Lynch, Ryan B. Choi, Tanmoy Roychowdhury, Renae L. Judy, Kaavya Paruchuri, Dong-Chuan Go, Sharika Bamezai, John Cabot, Sabina Sorondo, Michael G. Levin, Dianna M. Milewicz, Cristen J. Willer, Pradeep Natarajan, Saiju Pyarajan, Kyong-Mi Chang, Scott Damrauer, Phil Tsao, Stephen Skirboll, Nicholas J. Leeper, Derek Klarin

## Abstract

**Background:** The genetic risk of intracranial aneurysm (IA) development has been ascribed largely to the genetic risk of smoking exposure and hypertension. However, the relationship of IA to other cardiovascular traits and the contribution of IA risk loci to aberrant gene programs within cerebrovascular cell types remains unclear.

**Methods:** We performed a genome-wide association study in the Million Veteran Program testing association of roughly 25 million DNA variants with unruptured IA (3,165 cases and 592,927 controls) in veterans of European, African, and Hispanic ancestries. This was meta-analyzed with publicly available summary statistics to yield a final cohort of 15,438 cases and 1,183,973 controls. Candidate causal genes were prioritized through expression quantitative trait loci colocalization, fine-mapping transcriptome wide association studies, and multi-trait colocalization. We constructed a cerebrovascular single nucleus RNA sequencing (snRNA-seq) dataset and integrated IA summary statistics to prioritize candidate causal cell types. We then constructed a polygenic risk score to identify patients at greater risk of developing IA.

**Results:** We identified five novel loci association with IA, increasing the number of known susceptibility loci to 22. At these susceptibility loci, we prioritized 16 candidate causal genes. Amongst other cardiovascular traits, we found a significant positive genetic correlation of IA with coronary artery disease and abdominal aortic aneurysm, and we identified 13 IA risk loci that colocalize with aneurysmal, atherosclerotic, and blood pressure traits. Integration of an IA gene set with human cerebrovascular snRNA-seq data revealed significant association with matrix-producing pericytes, smooth muscle cells (SMCs), and other mural subtypes. In addition, gene expression analysis revealed enrichment of several candidate genes within SMCs and pericytes. Finally, a high polygenic risk score (PRS) was significantly associated with IA across European (OR: 1.87, CI: 1.61-2.17, P = 8.8 × 10^−17^), African (OR: 1.62, CI: 1.19-2.15, P = 1.2 × 10^−3^), and Hispanic (OR: 2.28, CI: 1.47-3.38, P = 1.0 × 10^−4^) ancestries.

**Conclusion:** Here, we identify five novel loci associated with IA. Integration of summary statistics with cerebrovascular snRNA-seq reveals association of cell-types involved in matrix production. We constructed and validated a PRS that predicts IA, while controlling for demographic variables including smoking status, sex, and blood pressure. Taken together, our findings suggest that an intrinsic deficit in matrix production and vascular integrity may drive IA pathogenesis independent of systemic hypertension and smoking exposure.

## INTRODUCTION

Intracranial aneurysms (IAs) are dilations of the cerebral arterial vasculature. These aneurysms represent a major cause of cerebrovascular morbidity, and the most feared complication – aneurysm rupture - results in intracranial hemorrhage, carrying a mortality of 50%.^1^ While a minority of patients develop syndromic forms of IAs due to connective tissue disorders, such as Marfan Syndrome, Ehlers Danlos Syndrome, or adult dominant polycystic kidney disease (ADPKD), 80-90% of patients with IA have no known predisposing genetic syndrome or immediate family history.^2–4^ In these patients, IA risk, similar to that of many complex traits, appears to be mediated by a combination of polygenic inheritance and environmental factors. The only multi-ancestry genome wide association study (GWAS) to-date estimated a narrow-sense genetic liability of 21% through common single nucleotide polymorphisms (SNPs).^5^ Hypertension and smoking, two of the strongest clinical risk factors for development of IA, not only share substantial genetic overlap with IA but likely have a causal genetic role in IA development as demonstrated by Mendelian Randomization.^6^

Although the clinical risk factors such as age, hypertension, smoking, and female sex have been well documented, the underlying etiology of IA and its relationship to other related cardiovascular traits such as atherosclerosis or aortic aneurysm is not well understood. Epidemiological studies have described an increased likelihood of IA development in patients with aneurysms in other vascular beds, suggesting common genetic or environmental risk factors.^7–11^ In addition, recent work has suggested common pathways and drug targets with atherosclerosis and abdominal aortic aneurysms (AAA), including *PCSK9*.^12, 13^ Appreciation of the shared genetic architecture of IA with these cardiovascular traits may help reveal other shared therapeutic targets.

To this end, we sought to expand upon known IA genetic susceptibility loci by performing a GWAS leveraging genetic data of patients in the Million Veteran Program (MVP), a precision medicine effort by the Department of Veteran’s Affairs to study how genes affect human health and disease. This cohort is enriched for patients with vascular pathologies and is ethnically diverse with patients from Hispanic and African ancestries in addition to European ancestries.^13–15^ We then used a combination of expression quantitative trait locus (eQTL) colocalization, fine-mapping transcriptome-wide association study (TWAS), and multi-trait colocalization to prioritize candidate causal genes and variants. We also assessed the pleiotropy of IA susceptibility loci, the genetic correlation of IAs to other cardiovascular traits, and the putative causal cell types involved through analysis of human cerebrovascular single nucleus RNA-sequencing (sn-RNA seq) data. Finally, we developed and validated the utility of a polygenic risk score (PRS) controlling for concomitant clinical risk factors to predict IA diagnosis.

## METHODS

### Study Populations

We conducted a discovery genome-wide association study (GWAS) using DNA samples and phenotypic data from the MVP and FinnGen (DF8), a publicly available biobank of Finnish participants’ clinical and genetic data. In MVP, individuals aged 18 to over 100 years have been recruited from 63 Veterans Affairs (VA) Medical Centers across the United States. After quality control, we identified 2,280 participants of European, 627 of African, and 258 of Hispanic ancestry with IA and 592,927 controls without evidence of IA. Within FinnGen, we identified 1,519 participants with IA and 284,164 controls.

### Intracranial Aneurysm Phenotype Definitions

From the participants passing quality control in MVP, individuals were defined as having IA based on possessing at least one of the following ICD codes for unruptured intracranial aneurysms: ICD9 473.3 and/or ICD10 I67.1 in their electronic health record (EHR). Patients with a diagnosis code for Marfan, Ehlers Danlos, ADPKD, and traumatic subarachnoid hemorrhage were excluded (Supplemental Table 1). After exclusion, controls (individuals without IA) were defined as participants without an IA diagnosis code.

### PheWAS of IA Risk Variants

Multiple databases with publicly available summary statistics were used to assess phenotype associations with IA lead variants. The NHIGRI-EBI GWAS catalog is a publicly available online database comprising 60,071 summary statistics from a wide array of phenotypes.^16^ PhenoScanner v2 is a similar dataset comprising 65 billion associations of 150 million variants.^17,18^ Finally, the Cardiovascular Disease Knowledge Portal (CDKP) (https://cvd.hugeamp.org/) is a database part of the Common Metabolic Diseases Knowledge Portal (CMDKP) project, and contains 207 GWAS summary statistics for 157 anthropometric and cardiovascular traits and conditions. IA lead risk variants, defined as those with the lowest p-value in a megabase window identified in our GWAS analysis, were queried in each of these three databases separately to perform the PheWAS (Phenome-wide Association Study). Phenotypic associations related to lab values, family history, medication use, imaging variables, or self-reported diagnoses were excluded. After exclusion, each trait was re-coded into a broad phenotypic category.

### Causal IA Gene and Variant Identification

We prioritized candidate causal genes at each of the IA risk loci by collating evidence from the following: 1) Literature review: prior genetic, clinical, or functional studies of genes at the locus were reviewed, 2) Proximity: the closest gene to the lead risk variant, 3) eQTL query: cis-eQTLs from the Genotype-Tissue Expression Project (GTEx)^19^ aortic and tibial tissue datasets with significant association P < 5×10^−6^, 4) TWAS: results from FUSION^20^, a fine-mapping technique to identify causal genes in a transcriptome-wide association study (TWAS) using bulk RNA-seq data from post-mortem aortic and tibial tissue and IA meta-analysis summary statistics, 5) eQTL colocalization analysis: colocalizing eQTLs were determined using IA meta-analysis summary statistics and annotated GTEx eQTLs in tibial and aortic tissue (echolocatoR^21^), 6) genes with protein-altering variants in high linkage disequilibrium (R^2^ > 0.8) with the lead IA risk variant (LDLinkR)^22^, 7) Multi-trait colocalization: prioritization of colocalizing variants among at least 1 set of cardiovascular summary statistics (Hyprcoloc).^23^ Genes prioritized as causal (beyond *SOX17* where prior literature has established this as the likely causal gene) were identified based on having: 1) two prioritization strategies highlighting it as a likely causal gene *and* 2) plausible biological evidence for a role in IA pathogenesis based on prior genetic, clinical, or functional studies.

### LD Score Regression Analysis

Genetic correlation of cardiovascular traits was assayed using LDSC function in the *GenomicSEM* R package. We obtained and used summary statistics for CAD^24^, PAD^15^, SBP/DBP (http://www.nealelab.is/uk-biobank/), AAA^25^, TAAD^26^, along with descending and ascending aortic diameters.^27^ We approximated the LD structure from Europeans within the 1000 Genomes reference panel^28^, and set a Bonferroni corrected P < 0.0014 (0.05/36 pairwise comparisons) for statistical significance.

### Construction and Annotation of Human Cerebrovascular snRNA-seq Data

Using publicly available data,^29, 30^ we identified cerebrovascular snRNA-seq data from 21 healthy control patients without any clinical or histopathological evidence of neurological or cerebrovascular disease. We also included patients from the Religious Orders Study and Memory and Aging Project (ROSMAP) without any neurologic pathology as used in Garcia et al.^30^ Data from each sample were processed in R Studio with *Seurat* (4.3.0). After conversion to a single cell experiment (SCE), doublets and ambient RNA were removed. Cells within each sample were filtered based on the following parameters: 200 < nFeatureRNA < 5000; 200 < nCount_RNA < 20,000; and percent.mt <0.5%. Normalization was performed using SCTransform v2. Each processed sample was then carried forward for integration. The integrated data set was normalized and an integrated analysis was applied to all ∼140,000 cells. Level 1 annotation was performed by transferring cell type labels with a high prediction value from the Tabula Sapiens^31^ vasculature single-cell reference set and using canonical markers for vascular and cerebral cell types as described in Garcia et al. and Yang et al. After subsetting cerebrovascular cell types including, endothelial cells, mural cells, fibroblasts, and astrocytes, we performed Level 2 annotation of these cell types, again using markers described in Garcia et al. and Yang et al. Differential expression of candidate causal genes was assayed using the ‘SCT’ assay.

*Cell Type Prioritization of IA Using Integrated snRNA-seq Data and IA Summary Statistics* A customized gene set using Stage 2 summary statistics effect sizes was created using MAGMA. The integrated cerebrovascular snRNA-seq dataset with level 2 annotations was converted to Anndata format for use with scanPy in the scDRS pipeline (https://github.com/martinjzhang/scDRS).^32^ We performed individual cell-level analysis to identify populations of cells with enrichment of putative candidate causal genes. Next, cell type-level analysis was performed to identify clusters within the level 2 annotation that are significantly associated with IA. Finally, gene-level analysis was performed to identify genes and gene programs correlated with the scDRS.

### Generation of an IA Polygenic Risk Score

A weighted polygenic risk score (PRS) was constructed using PRScs, which generates posterior genetic variant effect sizes throughout the genome using continuous shrinkage priors. Using effect sizes (beta) from Bakker et al. Stage 2 summary statistics, we created scores for 756,520 variants, estimated LD structure using the 1000 genomes European reference panel. We applied this score to MVP participants from European, African, and Hispanic ancestries. Each individual was additively scored based on copy number of scored risk alleles using Plink-score flag.

### Statistical Analysis

In our primary discovery analysis, genotyped and imputed DNA sequence variants in individuals of European, African, and Hispanic ancestry were tested for association with IA using logistic mixed models as performed in the REGENIE v2.0 statistical software program.^33^ We included in step 1 of REGENIE (i.e. prediction of individual trait values based on the genetic data) variants that were directly genotyped, had a minor allele frequency >1%, <10% missingness, and with Hardy-Weinberg equilibrium test P-value>10^−15^. The association model used in step 2 of REGENIE included as covariates age, sex, and 5 principal components of ancestry. Next, associated statistics across of European, African, and Hispanic ancestry MVP participants were meta-analyzed along with summary statistics available from FinnGen using an inverse variance-weighted fixed effects method as implemented in the METAL software program (Stage 1).^34^

We next performed meta-analysis with IA summary statistics from Bakker et al (Stage 2). Within the final meta-analysis, we defined IA susceptibility loci as those whose lead variant had nominal significance (p < 0.05) in both Stage 1 and Bakker et al. summary statistics, were directionally consistent across cohorts, and had an P < 5×10^−8^ in the meta-analysis. Novel loci were annotated if greater than 500 kilobases from a previously described IA susceptibility locus. All logistic regression values of P were two-sided.

In the PheWAS analysis, IA lead risk variants were queried using PhenoScanner v2, GWAS catalog, and cardiovascular disease knowledge portal, which are all online compilations of summary statistics from prior GWAS. We used a p-value threshold of P < 5×10^−8^ to determine significance (i.e. only phenotypes with variants at genome-wide significance were considered).

Genetic correlation of IA with cardiovascular traits was performed using LD score regression. Associations were considered significant if a Bonferroni corrected two-sided p-value was less than 0.0014 (.05/36 pairwise comparisons). Multi-trait colocalization was performed with Hyprcoloc, a deterministic Bayesian divisive clustering algorithms that can assess multiple traits simultaneously. Colocalizing traits with a posterior probability > 0.7 were considered significant. Similarly, significance of eQTL colocalization performed with echolocatoR, which also uses a Bayesian approach based on priors, was declared for genes with posterior probability > 0.7.

In our snRNA-seq analysis, cell-type level analysis was performed with scDRS. Benjamini-Hochberg corrected p-values were calculated and significant associations were declared with P-values < 0.05.

In our PRS analysis, logistic regression models were used to estimate odds ratios (OR) and 95% confidence intervals for associations for prevalent IA adjusting for age, sex, diastolic blood pressure, smoking status, and 5 principal components. Patients with diastolic blood pressures outside a range of 50-109, corresponding to the bottom and top 0.5 percentiles, were excluded. We calculated the OR of IA using the PRS as a continuous independent variable (standard deviation unit) along with a high PRS (top 5^th^ percentile of the continuous PRS). Bonferroni corrected P-values < 0.017 (0.05/3 ancestries) were deemed significant.

## RESULTS

### Multi-ancestry IA GWAS

We performed a two-stage GWAS (Figure 1). In Stage 1, we performed a logistic mixed model regression testing the association of TOPMED imputed genetic variants for each ancestry in the MVP. We obtained 15.4 million (African), 10.1 million (Hispanic), 8.9 million (European) common variants (MAF > 0.01) for analysis (Table 1). The resulting summary statistics were then meta-analyzed, also including summary statistics from a Finnish cohort study (FinnGen) comprising 13.0 million variants. The meta-analysis was comprised of 4,684 IA cases (2,280 European, 627 Black, 258 Hispanic, and 1,519 Finnish cases) along with 877,091 controls. We identified four loci at genome-wide significance, all of which have been previously reported (Supplemental Table 2).^5^ There was no evidence of genomic inflation in any group (Supplemental Figure 1). In addition, we replicated the 16/17 known loci with P < 0.05 in our Stage 1 analysis (Supplemental Table 3). In Stage 2, we meta-analyzed the results of Stage 1 with summary statistics of the prior IA GWAS (Bakker et al.) consisting of 10,754 cases and 306,882 controls from European and East Asian ancestries to yield a total of 15,438 cases and 1.2 million controls.

**Figure 1:**
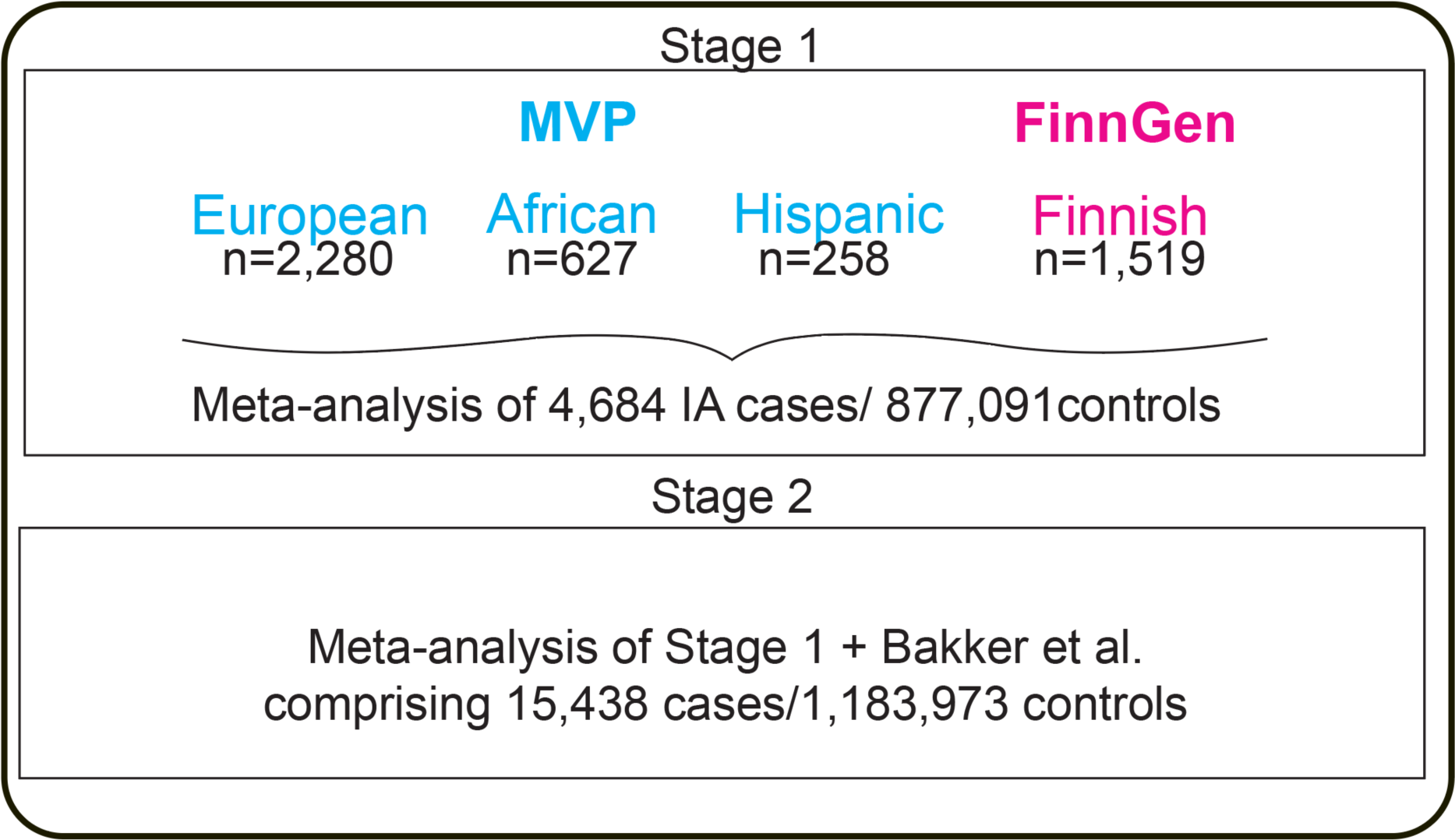
Intracranial Aneurysm GWAS Study Design. Stage 1: Meta-analysis of IA genome-wide associations across European, African, and Hispanic ancestries from MVP along with a Finnish cohort from FinnGen. Stage 2: Meta-analysis of Stage 1 with summary statistics from a prior intracranial aneurysm GWAS, to comprise 15,438 IA cases.

**Table 1:**
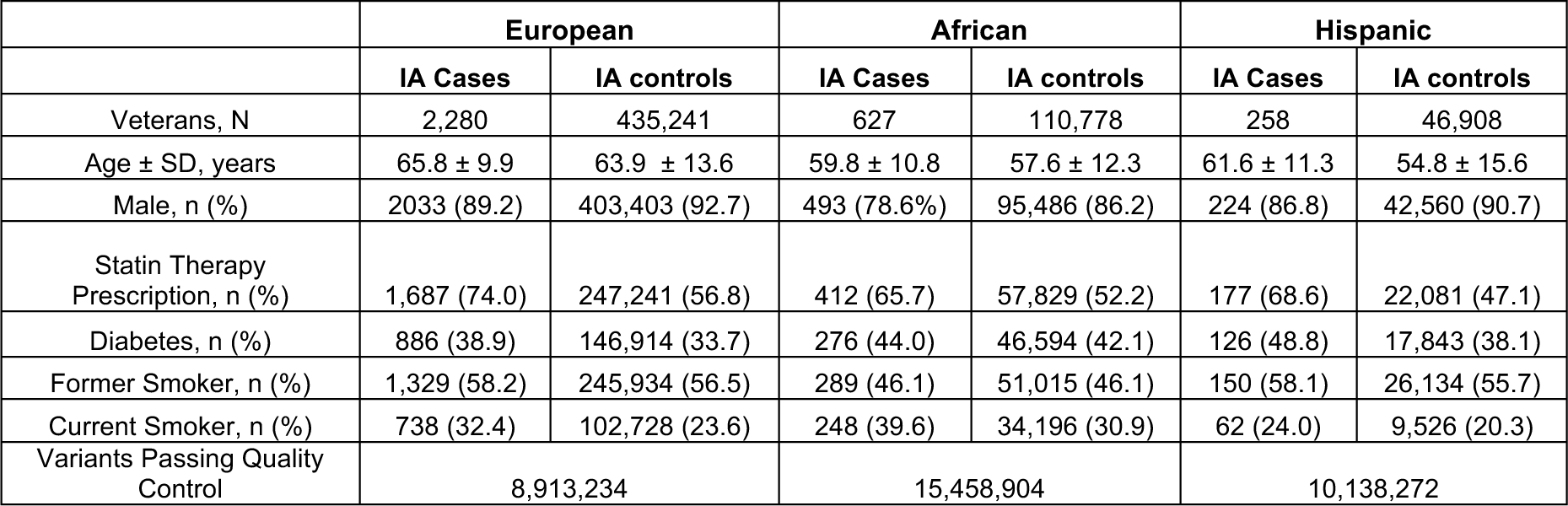
Demographic and Clinical Characteristics for Veterans in the MVP Discovery IA GWAS Analysis.

We identified 1,409 common variants (MAF > 0.01) at 22 loci associated with IA at genome wide significance (P < 5e-8), five of which were novel associations (Table 2). With the exception of the known lead variant near *BET1L* on chromosome 11, lead variants at the remaining 21 loci had at least nominal significance (P <0.05) in both Stage 1 and Bakker et al. summary statistics (Supplemental Table 3).^5^

**Table 2:** Five Novel IA Susceptibility Loci Identified in Stage 2.

| rsID | Chr | Locus | Consequence | EA | NEA | EAF | Effect (beta) | StdErr | Stage 1 P-value | Bakker et al. P-value | Stage 2 P-value |
| --- | --- | --- | --- | --- | --- | --- | --- | --- | --- | --- | --- |
| rs527725 | 1 | NAV1 | intron variant | a | c | 0.631 | 0.0771 | 0.0138 | 1.84E-05 | 1.86E-04 | 2.25E-08 |
| rs11099097 | 4 | FGF5 | intergenic variant | t | c | 0.313 | 0.0766 | 0.0138 | 1.10E-02 | 4.73E-07 | 2.98E-08 |
| rs59152651 | 6 | EXOC2 | downstream gene variant | a | g | 0.74 | -0.0965 | 0.0155 | 2.44E-05 | 4.23E-06 | 4.58E-10 |
| rs57301765 | 7 | TWIST1 | intergenic variant | a | g | 0.242 | -0.0967 | 0.0161 | 5.80E-04 | 9.21E-07 | 1.95E-09 |
| rs3118912 | 13 | DLEU1 | downstream gene variant | t | c | 0.215 | 0.094 | 0.0169 | 5.31E-05 | 1.20E-04 | 2.83E-08 |

Abbreviations: Chr, chromosome; EA, Effect Allele; NEA, Non Effect Allele; EAF, Effect Allele Frequency

Consistent with prior results, the lead variant in Stage 2 was rs1537373 (52.3% frequency of the G allele; OR = 1.20; 95% CI= 1.19-1.21; P = 8.7×10^−46^) within *CDKN2B-AS1* which encodes for the long non-coding RNA (lncRNA) *ANRIL*, associated with several cardiovascular and neoplastic disease states. We identified novel associations near *NAV1, FGF5, EXOC2, TWIST1,* and the lncRNA *DLEU1* (Supplemental Figure 2).

### Phenome-Wide Association Study

Pleiotropy of susceptibility variants offers insight into the shared pathophysiology of both similar and seemingly unrelated traits. To investigate the breadth of phenotypes associated with intracranial aneurysms, we performed a Phenome-wide Association Study (PheWAS) of IA susceptibility lead variants using publicly available summary statistics within Phenoscanner v2, NHGRI-EBI GWAS catalog, and the cardiovascular disease knowledge portal (CVDKP). We identified 316 unique associations of IA susceptibility loci over 12 broad phenotypic categories (Supplemental Table 4). A representative heatmap of traits with multiple associations and publicly available summary statistics is shown in Figure 2. As previously described, many of the IA susceptibility variants were also significantly associated with systolic and diastolic blood pressure. We also observed associations with coronary and peripheral arterial disease (CAD/PAD) as well as thoracic and abdominal aortic aneurysm. The lead variants at several loci including *MTMR3, STARD13, SOX17,* and *EXOC2*, were not significantly associated with any other cardiovascular traits, suggesting a unique association with IA. Surprisingly, at 7 loci including *TWIST1* and *EDNRA*, the IA risk variant was either associated with *decreased* SBP/DBP and or was *protective* for the development of CAD and PAD.

**Figure 2:**
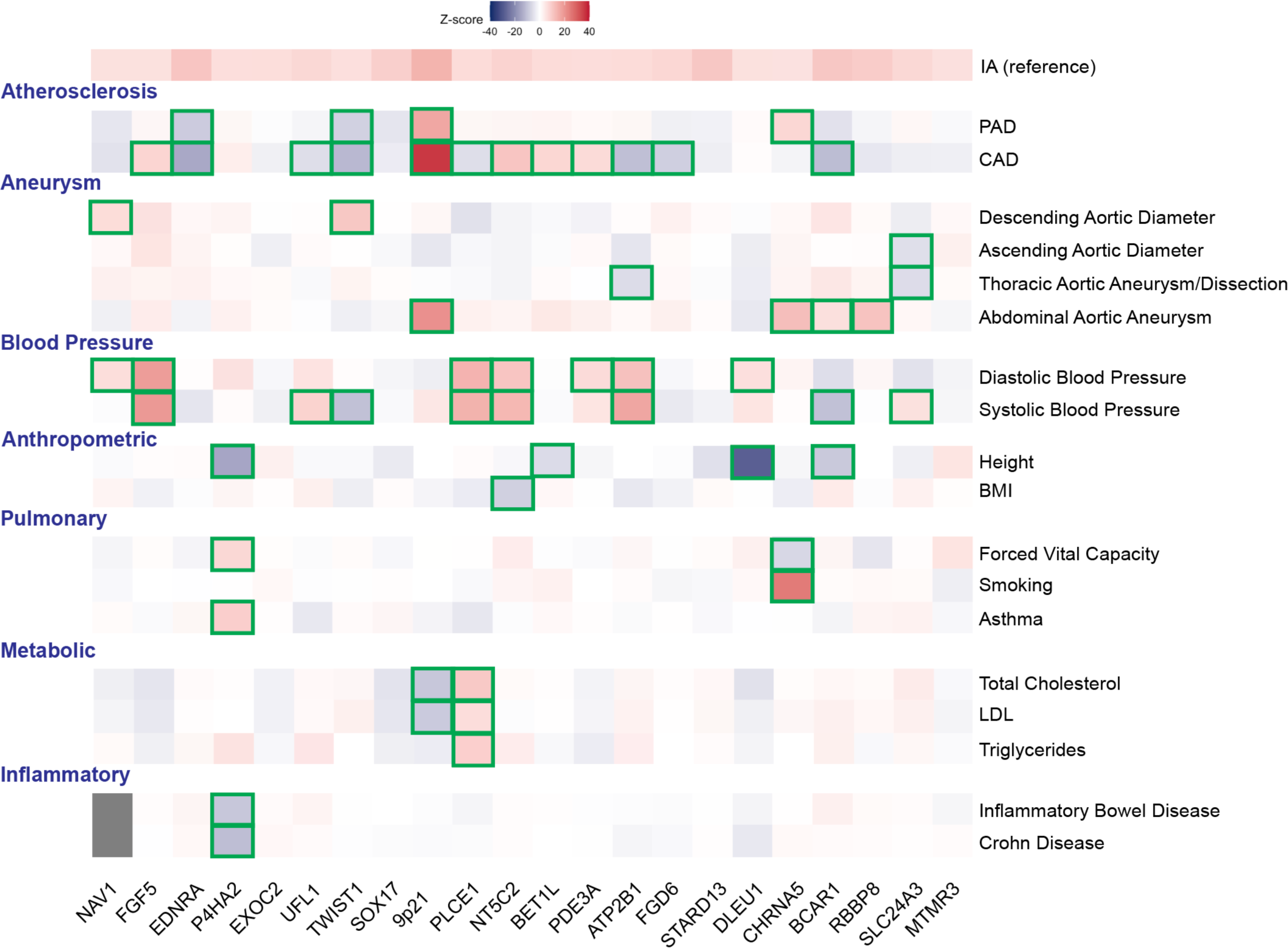
Representative Heatmap of Phenome-wide Association Results and Biologic Pathways Underlying IA Susceptibility Loci. Logistic regression Z-scores (aligned to the IA risk allele) from summary statistics of traits identified in PheWAS results at each of the 22 IA susceptibility loci. A positive Z-score (red) indicates a positive association between the PAD risk allele and the disease, whereas a negative Z-score (blue) indicates an inverse association. Boxes are outlined in green if the variant is associated with the indicated disease at genome-wide significance (logistic regression two-sided P < 5.0 × 10^−8^). IA, intracranial aneurysm; PAD, peripheral arterial disease; CAD, coronary artery disease; BMI, body mass index; LDL, low-density lipoprotein.

### Genetic Correlation of IA with Cardiovascular Traits

Given the extensive pleiotropy of IA risk loci with cardiovascular traits, we sought to assess the genetic correlation of IA with atherosclerotic, blood pressure, and aortic aneurysmal traits. We assessed the pair-wise genetic correlation of 9 cardiovascular traits using LD score regression (Figure 3a, Supplemental Table 5). Unsupervised clustering of traits based on genetic correlation coefficients (R_g_) revealed that IA shared heritability with aortic aneurysms, most significantly with AAA (R_g_ = 0.34, p = 2.9 × 10^−18^). In addition, similar to AAA, IA had a significant and positive genetic correlation with CAD (R_g_ = 0.17, p = 5.5 × 10^−6^).

**Figure 3:**
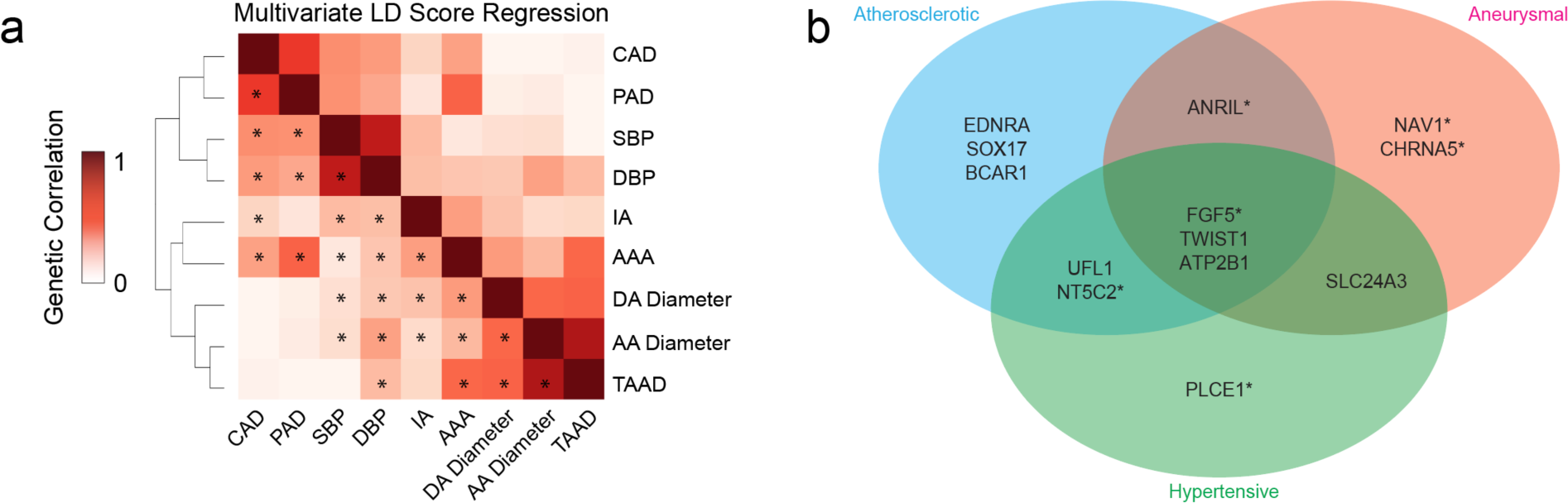
Genetic Correlation of Intracranial Aneurysm and other Cardiovascular Traits. a) Heatmap of pair-wise LD score regression across 9 cardiovascular traits. Dendrogram represents results of unsupervised clustering of traits based on correlation coefficient (R_g_). * denotes p<0.001. b) Venn-diagram illustrating colocalization of IA susceptibility loci with at least one trait categorized as atherosclerotic (CAD/PAD), hypertensive (SBP/DBP), or aneurysmal (AAA, DA size, AA size, or TAAD). * indicates significant colocalization (PP > 0.7) and concordant z-score direction of the lead variant.

While PheWAS can help identify shared phenotypic associations with lead variants and suggest common underlying biological pathways, it fails to assess whether shared genetic signals are likely driven by a common, causal variant. Therefore, we used multi-trait colocalization (Hyprcoloc) to determine the contribution of IA susceptibility loci to the observed positive correlation with atherosclerotic, blood pressure, and aneurysmal traits as seen by LD score regression (Figure 3b).^23^ We identified significant colocalization of *FGF5, TWIST1,* and *ATP2B1* with at least one trait within every category (posterior probability > 0.7). Notably, among the 9 IA susceptibility loci significantly colocalizing with at least one atherosclerotic trait, 6 prioritized variants had discordant effect directions with CAD. Similarly, 4 of the 6 IA susceptibility loci colocalizing with SBP or DBP are associated with reduced blood pressure. Thus, despite an overarching positive genetic correlation with CAD and SBP/DBP, it appears that certain prominent pathways that cause IA are paradoxically protective for hypertension and atherosclerosis.

### Prioritization of Candidate Genes

Prior human genetic and functional evidence strongly suggests that *SOX17* is a causal gene at one of our identified loci.^35, 36^ For genes at the remaining loci, we examined the literature for genetic or functional evidence of vascular involvement and identified non-synonymous coding variants in high LD with the lead variant (R^2^ > 0.8). In addition, we hypothesized that IA risk variants may be acting by inducing expression changes in *cis* and performed fine-mapping TWAS and colocalization analyses using aortic and tibial expression quantitative trait locus (eQTL) data from the Genotype Tissue Expression Project (GTEx) (Supplemental Table 6, Supplemental Figure 3). We supplemented this with multi-trait colocalization. When combining the above strategies, we identified 17 additional putative causal genes - *FGF5* (4q21), *EDNRA* (4q31), *P4HA2/SLC22A5* (5q31)*, UFL1* (6q16), *TWIST1* (7p21), *NOC3L/PLCE1* (10q23), *NT5C2* (10q24), *BET1L* (11p15), *ATP2B1* (12q21), *FGD6* (12q22), *STARD13* (13q13), *CHRNA5* (15q25), *BCAR1* (16q23), *SLC24A3* (20p11), and *MTMR3* (22q12) (Supplemental Table 7).

Interestingly, we found that increased *TWIST1* expression is associated with a decreased risk of IA. *TWIST1* encodes a master transcription factor involved in the regulation of smooth muscle cell plasticity during atherogenesis.^37^ Increased TWIST1 has been shown to promote a synthetic and proliferative phenotype in smooth muscle cells. Using multi-trait colocalization, we prioritized rs2107595 as the candidate causal variant (Supplemental Figure 4). This variant lies in a DNAse hypersensitivity site, and directly modulates *TWIST1* expression.^37^ Taken together, these results suggest that increased *TWIST1* expression promotes SMC proliferation and extracellular matrix production, reducing IA risk potentially through stabilization of the vascular wall.

### Gene expression analysis facilitates identification of candidate causal cell types

Our candidate causal gene set and PheWAS observations highlight directional discordance of many IA susceptibility loci with blood pressure and atherosclerosis. Thus, rather than systemic blood pressure and smoking status as sole mediators of IA pathogenesis, we hypothesized that intrinsic aberrancies within the cerebrovasculature may directly contribute to IA pathogenesis. To this end we analyzed expression of IA susceptibility loci using human cerebrovasculature single-nuclear RNA sequencing (snRNA-seq) data.

Using publicly available human brain snRNA-seq datasets enriched for cerebrovasculature we identified 20 patients without any neurological or vascular pathology.^29, 30^ We performed quality control and integrated snRNA-seq data from these patients with data from ROSMAP (see methods) to obtain over 147,439 nuclei and 31 Louvain clusters. After unsupervised clustering, we performed a Level 1 annotation to identify fibroblast, pericyte, endothelial, and smooth muscle cells as well as cerebral cell types (Supplemental Figure 4).

After subsetting relevant cerebrovascular cell types (endothelial cells, smooth muscle cells, pericytes, astrocytes, and meningeal/perivascular fibroblasts) we obtained 102,140 nuclei for analysis. We performed Level 2 annotation, using previously described markers to annotate endothelial, pericyte, fibroblast, and astrocyte subtypes (Figure 4a). Analysis of subpopulations of EC revealed enrichment of capillary ECs relative to arterial and venous ECs (Supplemental Figure 6). Our integrated analysis allowed identification of a relatively sparse population of ECs expressing Laminin alpha-2 and Laminin beta-1, important constituents of the basement membrane and blood brain barrier (Supplemental Figure 7). Concordant with the original datasets, these cells are annotated as tip ECs. Mural cells demonstrated the greatest amount of heterogeneity with four subclusters described in the cerebrovasculature. Amongst the pericytes, we annotated roughly equal proportions of matrix producing pericytes (M-pericyte) and pericytes involved in membrane transport (T-pericyte) (Supplemental Figure 6). M-pericytes display enrichment of type 4 collagen, while T-pericytes are enriched in several solute carriers (Supplemental Figure 7). Finally, the majority of fibroblasts displayed a perivascular phenotype as evinced by expression of structural components, while meningeal fibroblasts expressed several SLC solute transporters (Supplemental Figure 6c).

**Figure 4:**
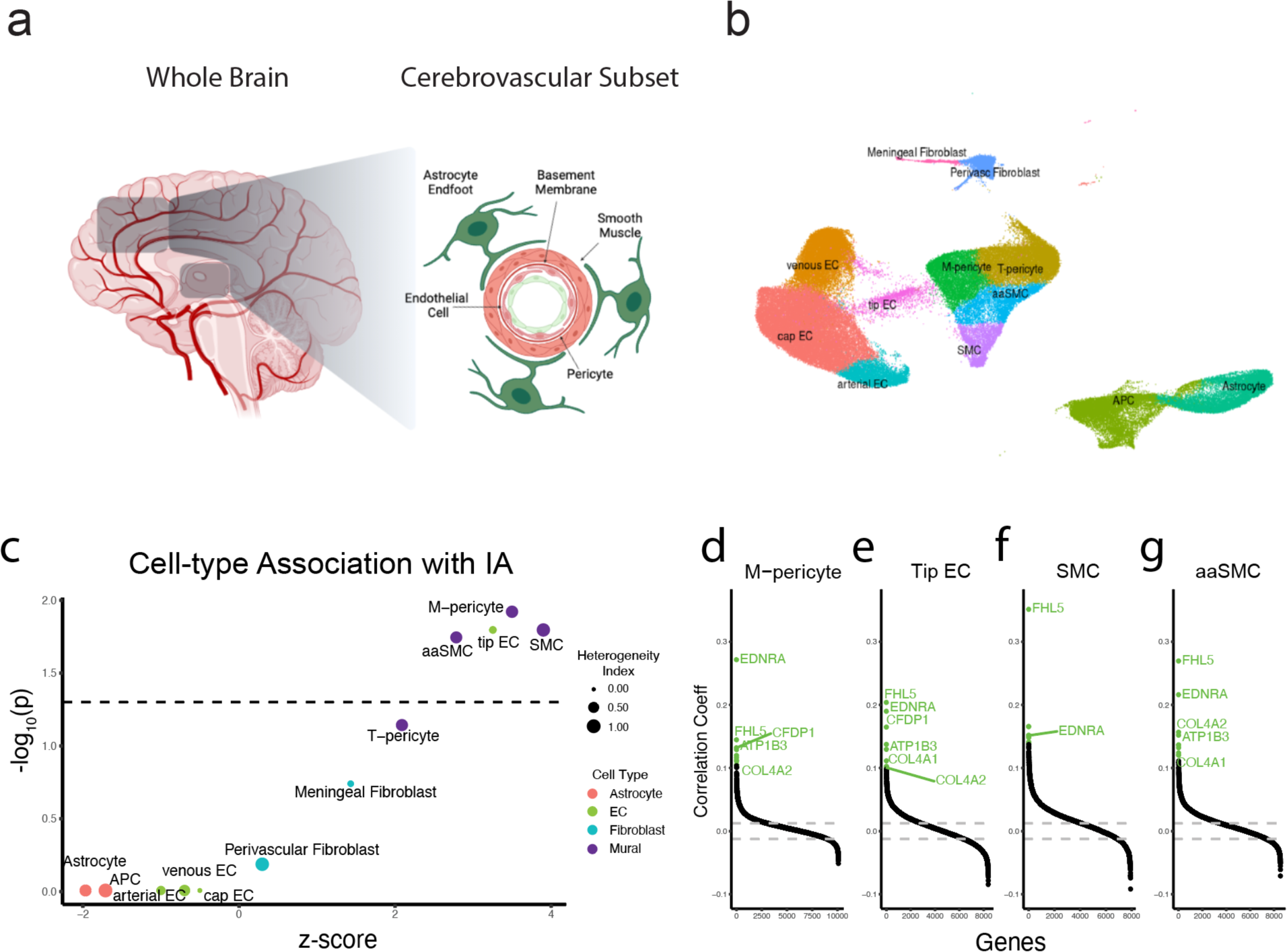
Generation of Human Cerebrovascular snRNA-seq Dataset and Integration with Intracranial Aneurysm Gene Set. a) Schematic depicting integration of healthy human cerebrovascular samples, Level 1 annotation of all cell-types, sub-setting of cerebrovascular cell types, followed by Level 2 annotation of cerebrovascular cell types. b) Cell-type level analysis depicting IA association with each Level 2 annotated cell-type. Cell-types are color-coded by their Level 1 annotation. Heterogeneity index reflects the degree of heterogeneity of the single cell disease relevance score in each cell-type. High heterogeneity suggests the presence of a subpopulation of cells with a high disease relevance score. c-f) Gene-level analysis illustrating correlation of each gene with disease relevance score across the mural cell types. EC, endothelial cell; NPC, neural progenitor cell; OPC, oligodendrocyte progenitor cell; APC, astrocyte progenitor cell; SMC, smooth muscle cell; aaSMC, arteriolar smooth muscle cell. Created with BioRender.com

Having defined the major and minor cell populations in the integrated snRNA-seq cerebrovascular dataset, we next sought to further prioritize candidate causal cell-types and gene programs through linking our GWAS and snRNA-seq datasets at single cell resolution. We integrated gene expression profiles with a gene set derived from IA summary statistics (MAGMA) to associate single cells with IA through computation of a single cell disease relevance score (scDRS).^32^ We began with a cell-type level analysis to identify cell type associations and heterogeneity within each cell type. SMCs, aaSMC, M-pericytes, and tip ECs were all significantly associated with IA (Figure 4b, Supplemental Table 8). With the exception of tip ECs, each cell type displayed considerable and significant heterogeneity suggesting subpopulations that contribute to the observed association. Single cell analysis of disease relevance score revealed a sub-population of M-pericytes and SMCs with the strongest signal (Supplemental Figure 7).

Among cell types significantly associated with the IA (mural cell populations and tip ECs), we assayed the correlation of each gene’s expression with the scDRS. We consistently found that *EDNRA* and *FHL5*, whose dysregulation was recently found to drive vascular calcification in atherosclerosis, were highly correlated with the scDRS. Both genes have high z-scores within the IA gene set and likely drive the observed cell-type association. Interestingly, within the M-pericyte, tip EC, and aaSMC populations we also found basement membrane components *COL4A1/COL4A2* as one of the most strongly correlated genes despite the lack of significant association signal within our GWAS summary statistics (Figure 4c-f, Supplemental Figure 8).

Finally, we sought to identify relevant cell types for the individual putative causal IA risk genes examining the cell-type enrichment of each candidate causal gene. Within ECs, we found enrichment of *BCAR1*, which encodes a scaffold protein involved in mechanotransduction.^38–40^ Within the mural cell types, we found enrichment of *EDNRA* and *PLCE1*, with relative enrichment of these genes in the M-pericyte population. Amongst the fibroblasts, *TWIST1* and *FGD6* demonstrated enrichment within both fibroblast populations, while *SLC24A3* expression was clearly enriched within the meningeal fibroblast population (Figure 5). Together, this suggests that although SMCs and pericytes appear to be the primary drivers of IA, dysregulation of ECs and fibroblasts may also contribute to IA development.

**Figure 5.**
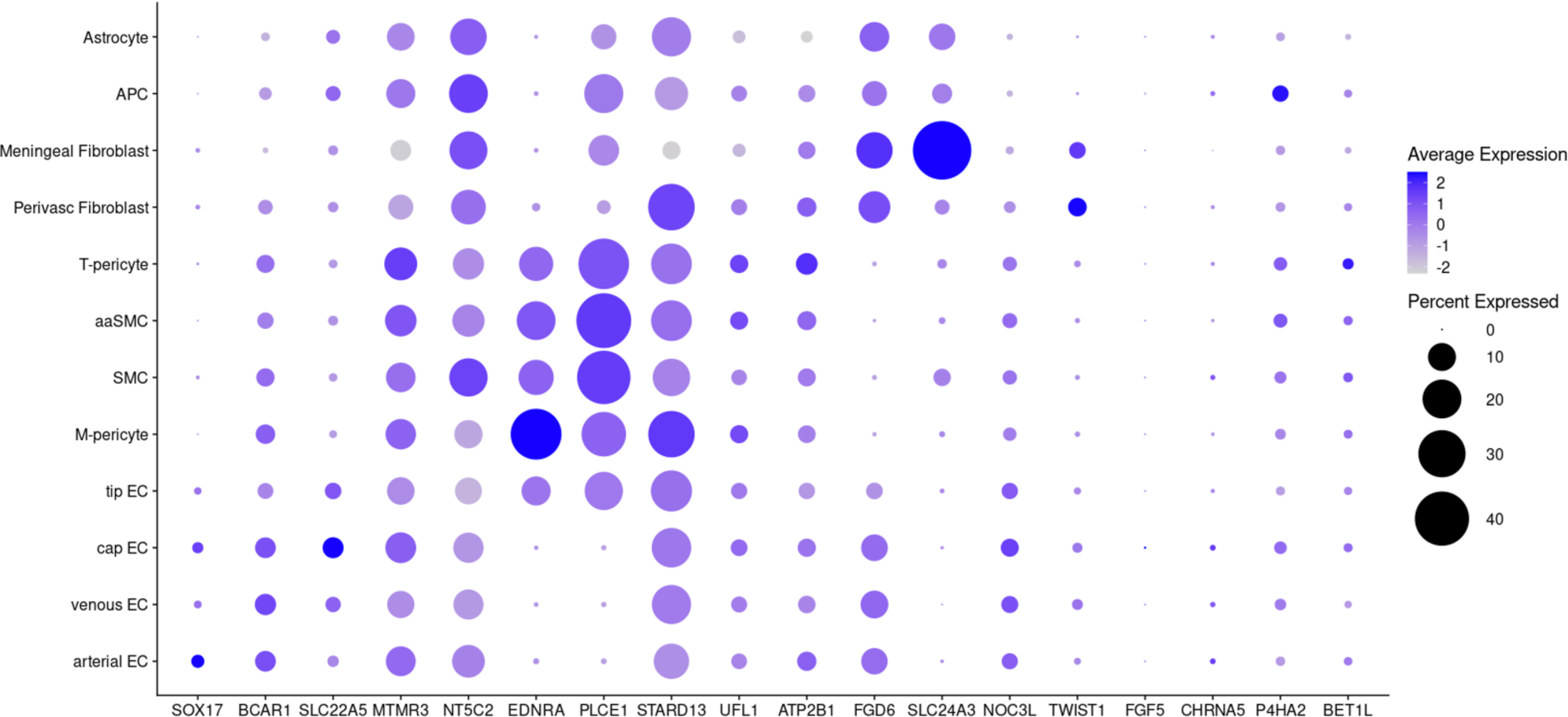
Gene Expression of Candidate Causal Genes within Cerebrovascular Cell Types. Dot-plot representing cell-type specific expression of our candidate causal gene set. Dot size represents proportion of cells with expression of the specified gene. Color gradient represents the average of normalized expression values with dark blue indicating high expression and light blue indicating low expression.

### Generation of an IA polygenic risk score

Finally, we sought to evaluate the contribution of polygenic inheritance on IA risk. Using the Bakker et al. summary statistics comprising over 10,000 cases, we generated a PRS using 756,520 variants (PRScs^41^, see methods). We assessed the performance of the PRS using patients within the MVP from European, African, and Hispanic ancestries separately. We identified patients with a high PRS (top 5^th^ percentile) who we hypothesized were most susceptible to IA development. Amongst the European cohort, patients in the highest ventile had an 87% increased risk of prevalent diagnosis (OR: 1.87, CI: 1.61-2.17, P = 8.82 × 10^−17^) while controlling for age, diastolic blood pressure, female sex, and smoking status in the regression model (Figure 6a). Similarly, patients with a high PRS with African and Hispanic ancestries show an increased risk of IA diagnosis (African: OR 1.62, CI 1.19-2.15, P= 1.18 × 10^−3^; Hispanic: OR 2.28, CI 1.47-3.38, P = 1.02 × 10^−4^) (Figure 6b-c). We found a similar effect while examining a 1-SD increase in PRS across ancestries. Finally, we stratified patients from each ancestry by ventile and observed an increasing prevalence and risk of IA, controlling for the aforementioned comorbidities and demographic parameters (Supplemental Figure 9).

**Figure 6.**
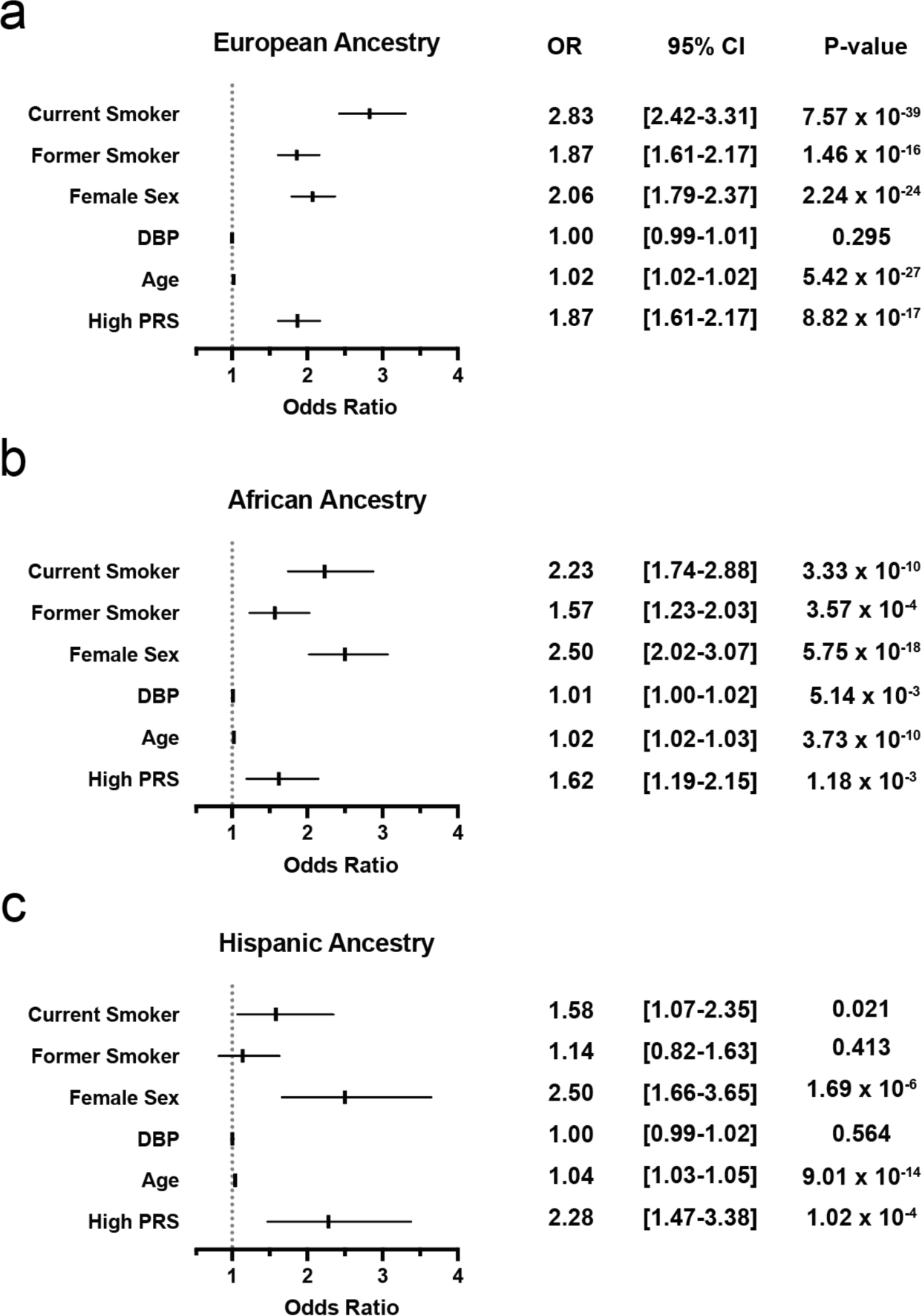
Validation of a Polygenic Risk Score to Predict Intracranial Aneurysm. Logistic regression results in patients within the MVP stratified by a) European ancestry b) African ancestry, and c) Hispanic ancestry. Forest plots (left) depict the odds ratio (OR) and 95% confidence intervals (95% CI) of ICA diagnosis in patients within the top 5^th^ percentile of a polygenic risk score, controlling for current/former smoking, sex, age, diastolic blood pressure (DBP) and the first five principal components.

## DISCUSSION

In this study, we performed a GWAS to identify 5 novel IA susceptibility loci, raising the total number of replicated IA loci to 22. As seen in our PheWAS, the vast majority of loci overlapped with susceptibility loci of several complex cardiovascular traits, namely atherosclerosis, blood pressure, and aneurysms. Consistently, we found a positive genetic correlation of IA with many cardiovascular traits and identified the causal genes contributing to the observed correlation by colocalizing signal from multiple sets of summary statistics. We identified causal cerebrovascular cell types by constructing a large snRNA-seq dataset from cerebrovascular tissue and integrating gene expression with IA summary statistics. Finally, we developed a polygenic risk score for IA and performed validation using the MVP.

These findings permit several conclusions. First, we show through several lines of evidence that IA has substantial shared heritability with other cardiovascular traits and hypothesize that this information may be useful for therapeutic intervention. Assessment of genetic correlations with LD score regression supports significant overlap with atherosclerosis, hypertension, and other aneurysmal traits. Of all cardiovascular traits, IA demonstrated the strongest genetic correlation with AAA, consistent with epidemiological and observational studies.^8, 9^ Furthermore, we applied fine-mapping with multi-trait colocalization to reveal the loci largely responsible for the observed correlation. As GWAS become increasingly powered and the number of significant associations with cardiovascular pathologies grows, assessing commonalities between phenotypes will be important to postulate shared mechanisms and therapeutic targets. Here, we find that the overlapping genetic correlation with atherosclerosis, aortic aneurysm, and hypertension is largely driven by six loci, 9p21 (atherosclerosis, aortic aneurysm), *NT5C2* (hypertension and atherosclerosis), *NAV1* (aortic aneurysm), *CHRNA5* (aortic aneurysm, smoking exposure^42^), *PLCE1* (hypertension), and *FGF5* (aortic aneurysm, atherosclerosis, hypertension). While further mechanistic validation of these loci remains to be performed, our data suggests the presence of a subset of shared, targetable pathways that may treat hypertension, aneurysmal disease, and atherosclerosis. In particular, the causal gene at the FGF5 locus (prioritized here to be *FGF5*) may be targeted to treat all three categories of cardiovascular pathology.

Second, despite an overall positive genetic correlation of IA with atherosclerosis and blood pressure traits, the presence of opposing risk alleles at multiple, shared genome-wide significant loci highlights the importance of vessel wall integrity in IA pathogenesis. In our PheWAS, the effect directions of the IA risk allele are protective for hypertension or CAD at several loci, including *EDNRA*. Increased expression of *EDNRA* is associated with atherosclerosis and hypertension through promotion of systemic vasoconstriction. A paradoxical *increased* risk of IA conferred by *EDNRA* variants associated with *decreased* blood pressure suggests an alternative mechanism by which this variant causes IA. *EDNRA* encodes a G-protein coupled receptor, which after binding its primary ligand endothelin-1, activates a plethora of signaling cascades involved in matrix deposition, contraction, and proliferation.^43^ The *EDN1*/*EDNRA* axis has both a critical role in neural crest specification, which ultimately gives rise to the craniofacial vasculature, and promotes extracellular matrix contraction which is likely to be critical in vessel wall integrity.^44, 45^ Similarly, variants at the *UFL1*-*FHL5* locus, also with discordant effect direction in the contexts of IA and CAD, have recently been shown to promote phenotypic switching of smooth muscle cells to a synthetic phenotype in the context of atherosclerosis.^37, 46^ Consistent with the concept that differences in matrix production can differentially drive atherosclerosis and IA, we find through integration of snRNA-seq and GWAS summary statistics that tip ECs and M-pericytes, both involved in basement membrane production through expression of type IV collagen and laminins, are significantly associated with IA. Moreover, amongst these cell types, *COL4A1*/*COL4A2* expression, which encode subunits of type IV collagen, the primary component of the basement membrane, is strongly correlated with the weighted expression of an IA gene set. Together these data suggest that shared causal pathways seen in IA and other cardiovascular traits may result in divergent phenotypic consequences. Therefore, despite considerable genetic overlap with atherosclerosis and hypertension, we hypothesize that *targeted* IA therapeutics augmenting extracellular matrix production and vascular wall stability will provide the most utility for IA treatment or prevention.

Third, our results suggest that focusing on SMC biology is likely to be particularly key for understanding IA pathologic mechanisms and identifying novel therapies. Through our genome-wide and gene-based transcriptomic analyses, SMCs and M-pericytes consistently emerged as likely causal cell types. Often referred to as ‘phenotype switching’, SMCs are capable of remarkable plasticity which typically involves downregulation of SMC markers and transitioning from a contractile phenotype to a proliferative, synthetic, or migratory phenotype.^47^ In the context of atherosclerosis, smooth muscle cells are thought to migrate to the intima and trans-differentiate to a fibrochondrocyte-like cell to secrete a collagen-rich ECM of the fibrous cap. In this manner, modulated SMCs can contribute to plaque burden. By contrast, in our study we annotated mediators of phenotype switching, including *TWIST1*, *EDNRA*, and *UFL1*, as likely to *reduce* risk of IA.^37, 48–50^ These genes likely promote vascular wall stability through ECM generation and smooth muscle cell proliferation. Consistent with this mechanism, inherited risk at *9p21* may cause SMC apoptosis and medial thinning through dysregulation of *CDKN2B*.^51^ Therefore, in the context of aneurysmal disease, harnessing the plasticity of SMCs to promote matrix production may restore vascular wall integrity and decrease the risk of aneurysm rupture. We also caution that emerging endeavors to generate therapeutics inhibiting SMC phenotype switching for atherosclerosis also consider potential deleterious effects of matrix destabilization in the context of concomitant aneurysmal degeneration.

Our findings should be interpreted within the context of the following limitations. First, our IA phenotype is based on EHR diagnosis code data, and may result in misclassification of case status. However, such misclassification should, on average, reduce statistical power for discovery and bias results toward the null. Second, with available ICD codes we were unable to ascertain patients who presented with intracranial hemorrhage from an IA, which limits our ability to identify risk loci for rupture. Third, the Veterans Affairs Healthcare System population is overwhelmingly male and our ability to detect sex-specific genetic associations in discovery was limited.^45^ Finally, although MVP participants with the highest PRS are at increased risk for IA, the PRS mechanism of action represents a combination of many causal risk factors, rather than a single pathway that leads to disease. Assessment of individual risk can aid in identifying those at highest risk for IA and more likely to obtain benefit from screening, regardless of mechanism.

In conclusion, we have performed the largest IA GWAS study to date and discovered 5 novel loci. We identify causal risk factors, cell types, and genes that may be used to inform clinical care.

## Data Availability

The full summary level association data from the MVP IA discovery analysis and meta-analysis from this manuscript will be available through dbGAP, accession code phs00167.v2.p1 upon publication.

---

**Supplemental Figure 1:**
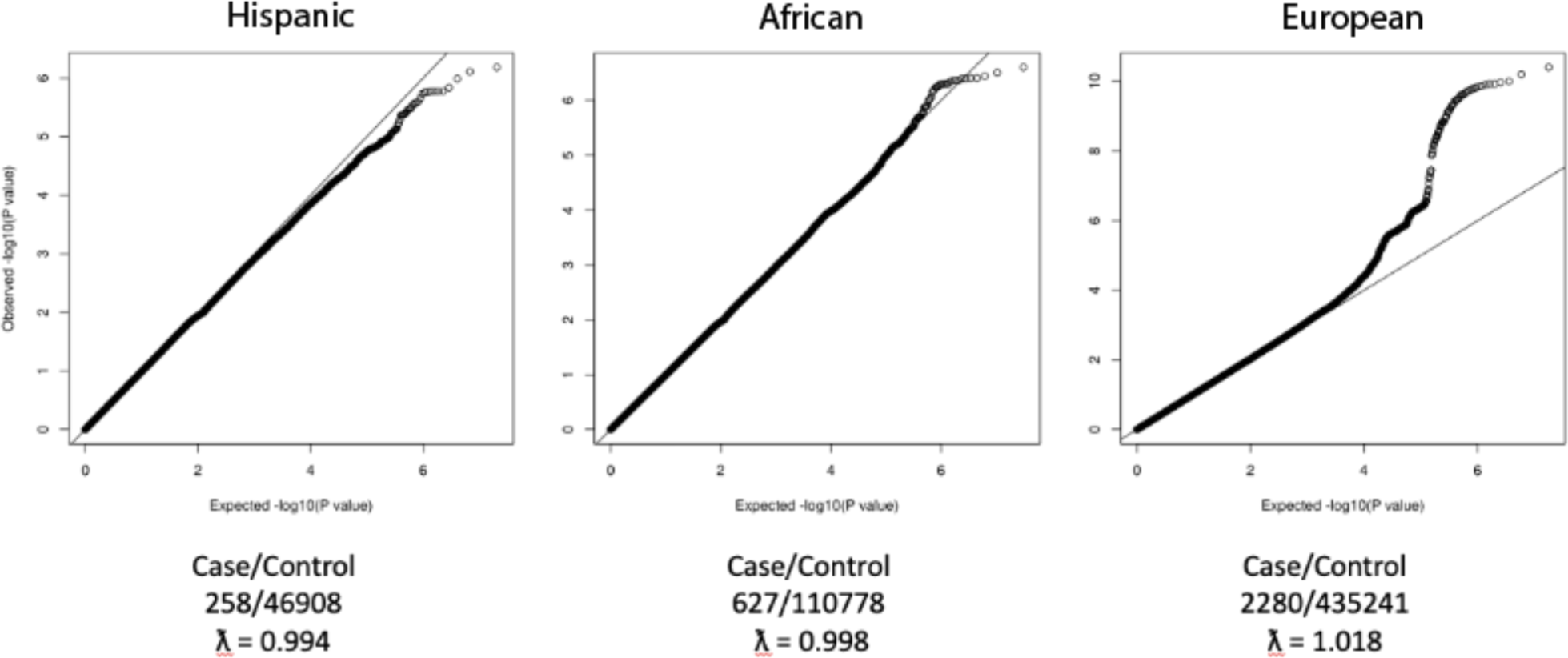
Quantile-Quantile (QQ) plots depicting the expected logistic regression association P-values versus the observed distribution of P-values for GWAS of each MVP ancestry. No systemic inflation was observed. All P-values were two-sided. GWAS, Genome-wide Association Study; MVP, Million Veterans Program

**Supplemental Figure 2:**
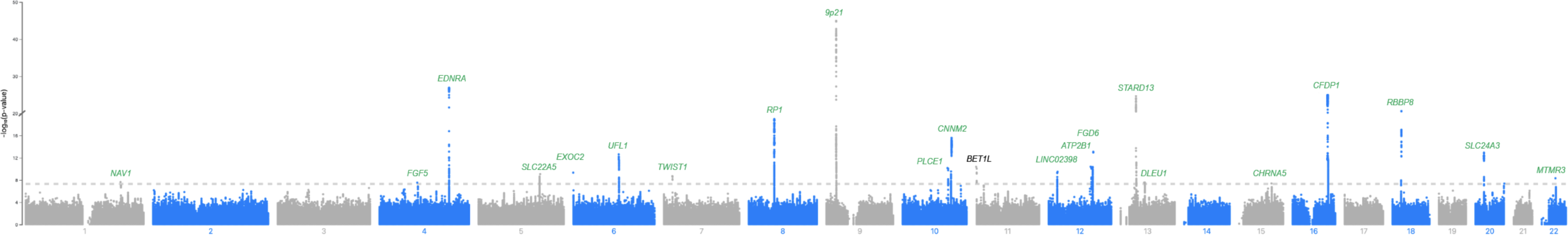
Manhattan plot of Stage 2 summary statistics depicting association of genotyped and imputed variants with intracranial aneurysm. Dashed line indicates genome-wide significance threshold of p < 5×10^−8^. Annotated genes represent closest gene to lead variant. Green text denotes loci replicated in Bakker et al. summary statistics with at least nominal significance (p < 0.05)

**Supplemental Figure 3:**
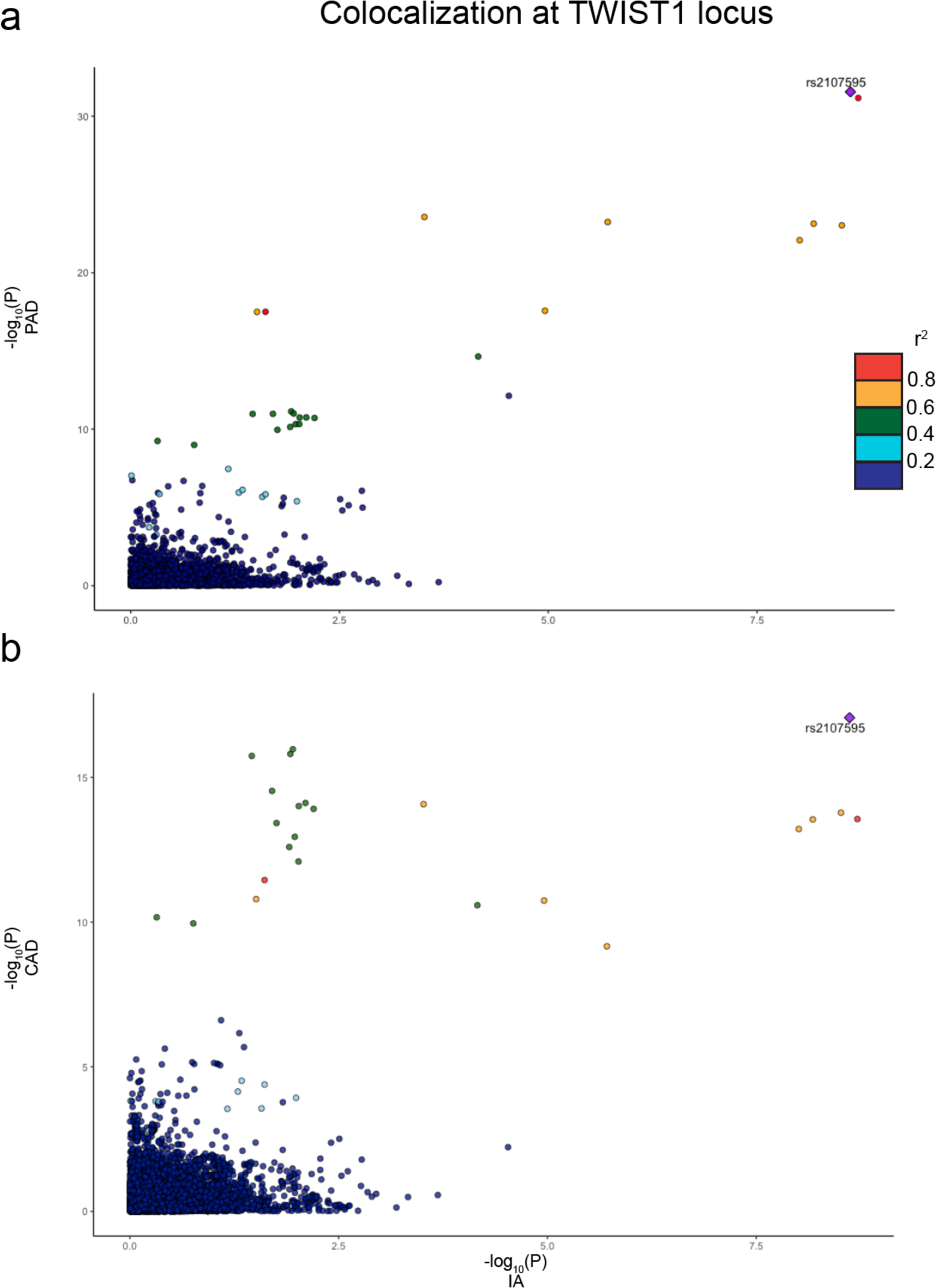
Comparison of association P-values at TWIST1 locus for IA (x-axis) and (a) CAD and b (PAD). Individual genetic variants are represented as points, color coded by degree of genetic correlation (r^2^) with the prioritized colocalizing variant (diamond).

**Supplemental Figure 4:**
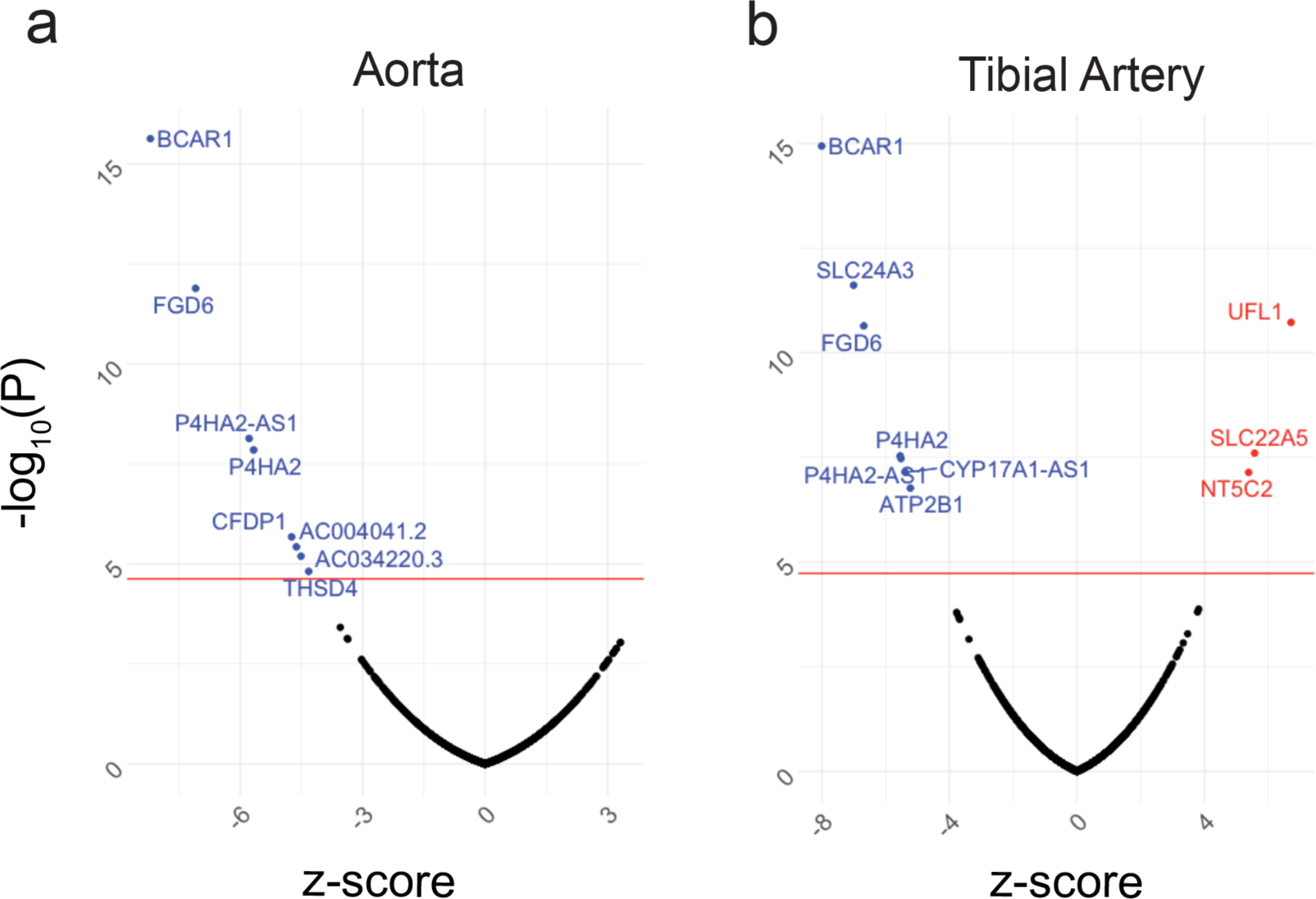
Fine mapping transcriptome-wide association study using bulk RNA-sequencing data from (a) aortic and (b) tibial tissue.

**Supplemental Figure 5:**
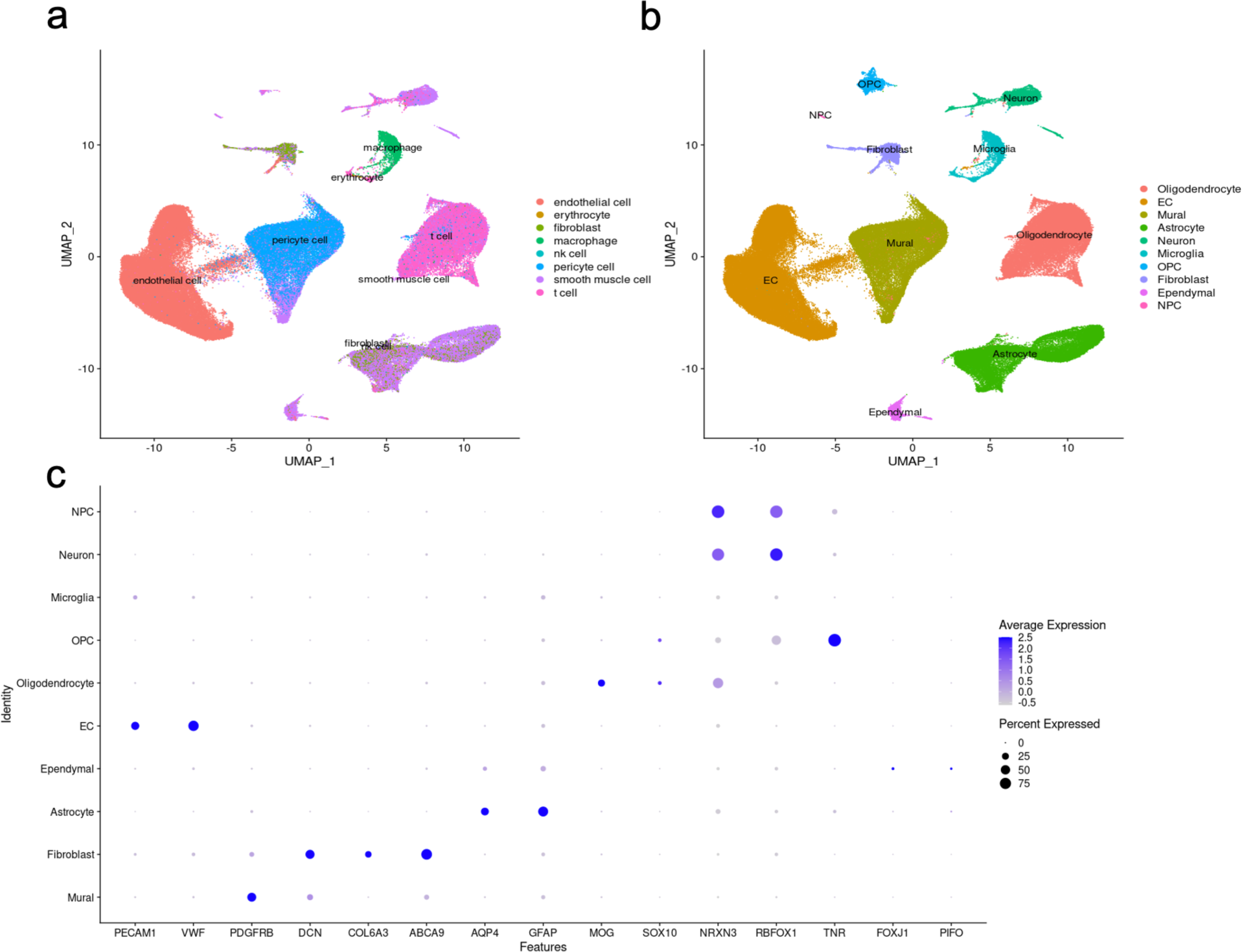
Level 1 annotation of brain cerebrovasculature. a) UMAP representation of predicted vascular cell types using Tabula Sapiens reference annotation. b) Final level 1 cell type annotation using Tabula Sapiens and c) dot plot representation of canonical marker expression for each cell type.

**Supplemental Figure 6:**
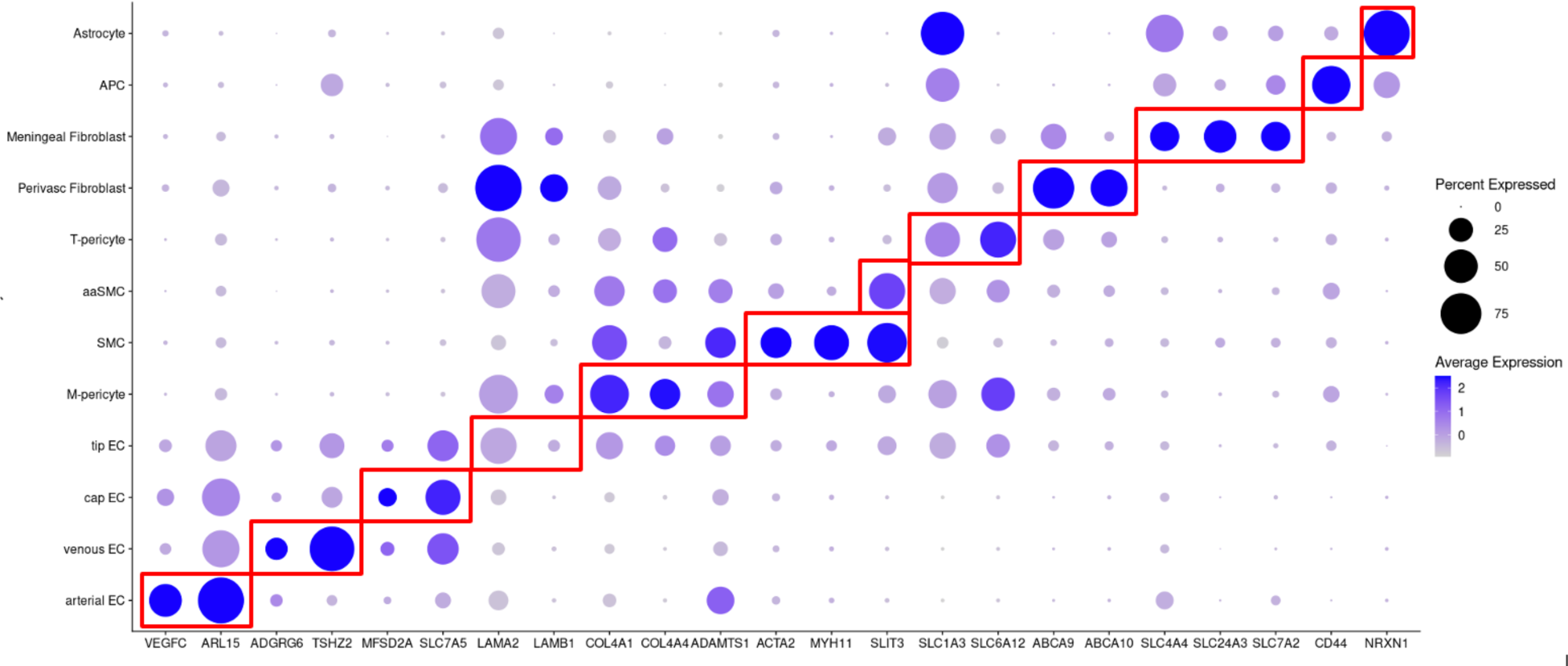
Level 2 Annotation of cerebrovasculature using previously described markers of vascular subtypes. Dot plot representing gene expression on vascular subtypes. Subtypes were annotated based on expression of genes highlighted in red boxes

**Supplemental Figure 7:**
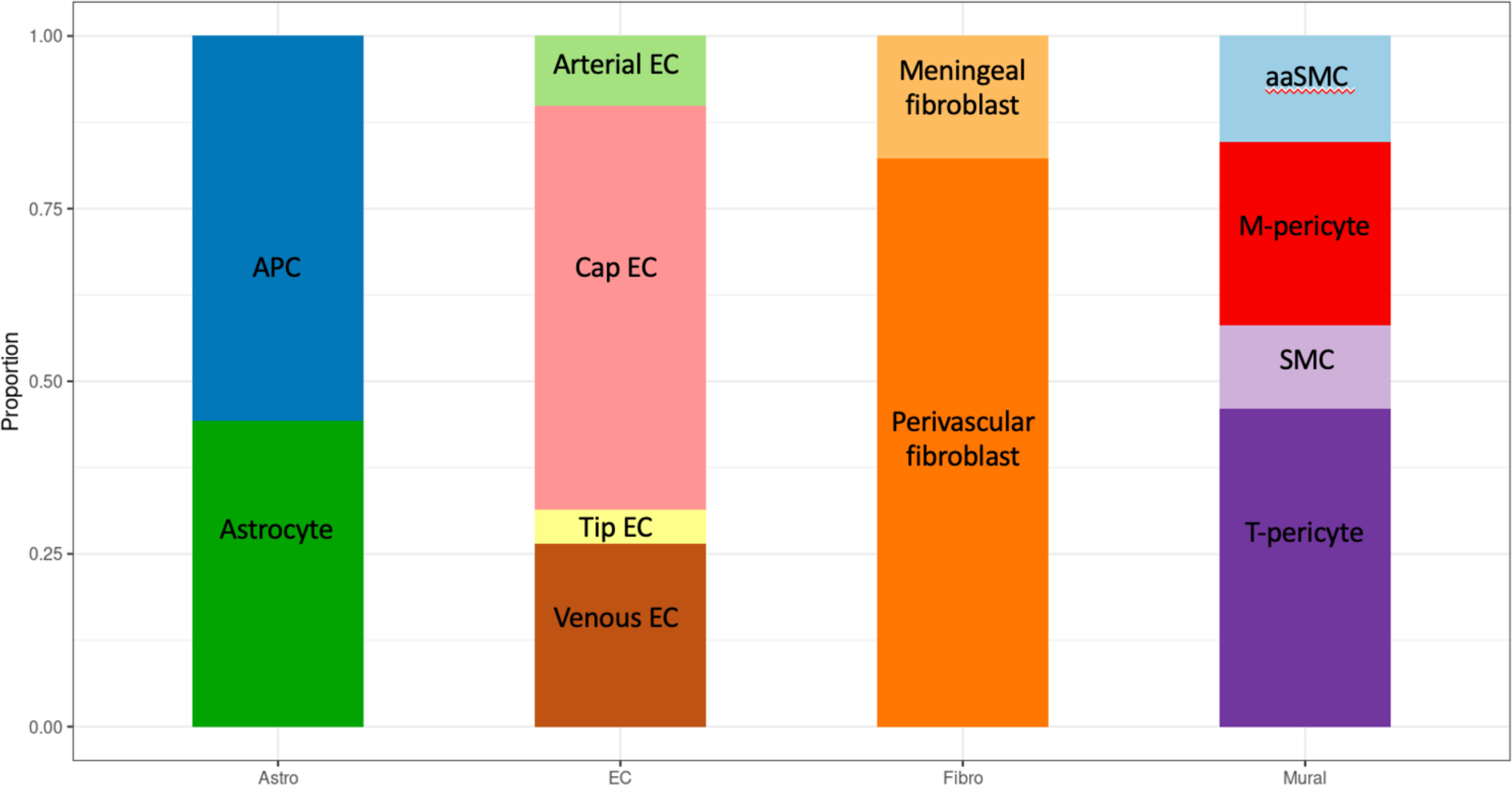
Proportions of cell sub-types amongst vascular stromal cell populations. aaSMC, arteriolar SMC; Cap EC, capillary EC; M-pericyte, matrix-pericyte; T-perictye, transport-pericyte

**Supplemental Figure 8:**
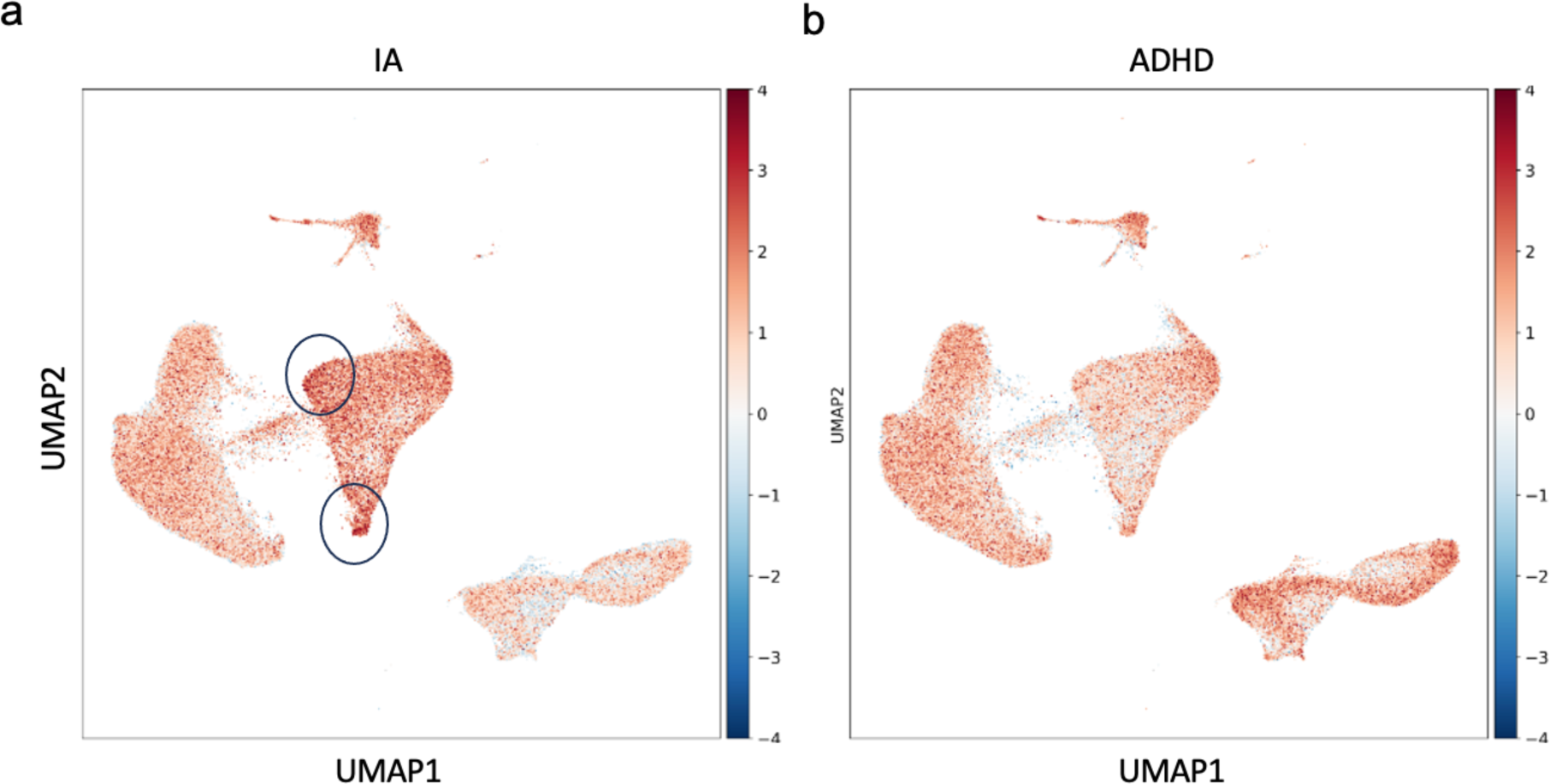
Single cell resolution of single cell disease relevance scores using a) Intracranial Aneurysm (IA) gene set and b) Attention deficit hyperactivity disorder (ADHD) gene set as a control. Cells with high scDRS scores highlighted in (a).

**Supplemental Figure 9:**
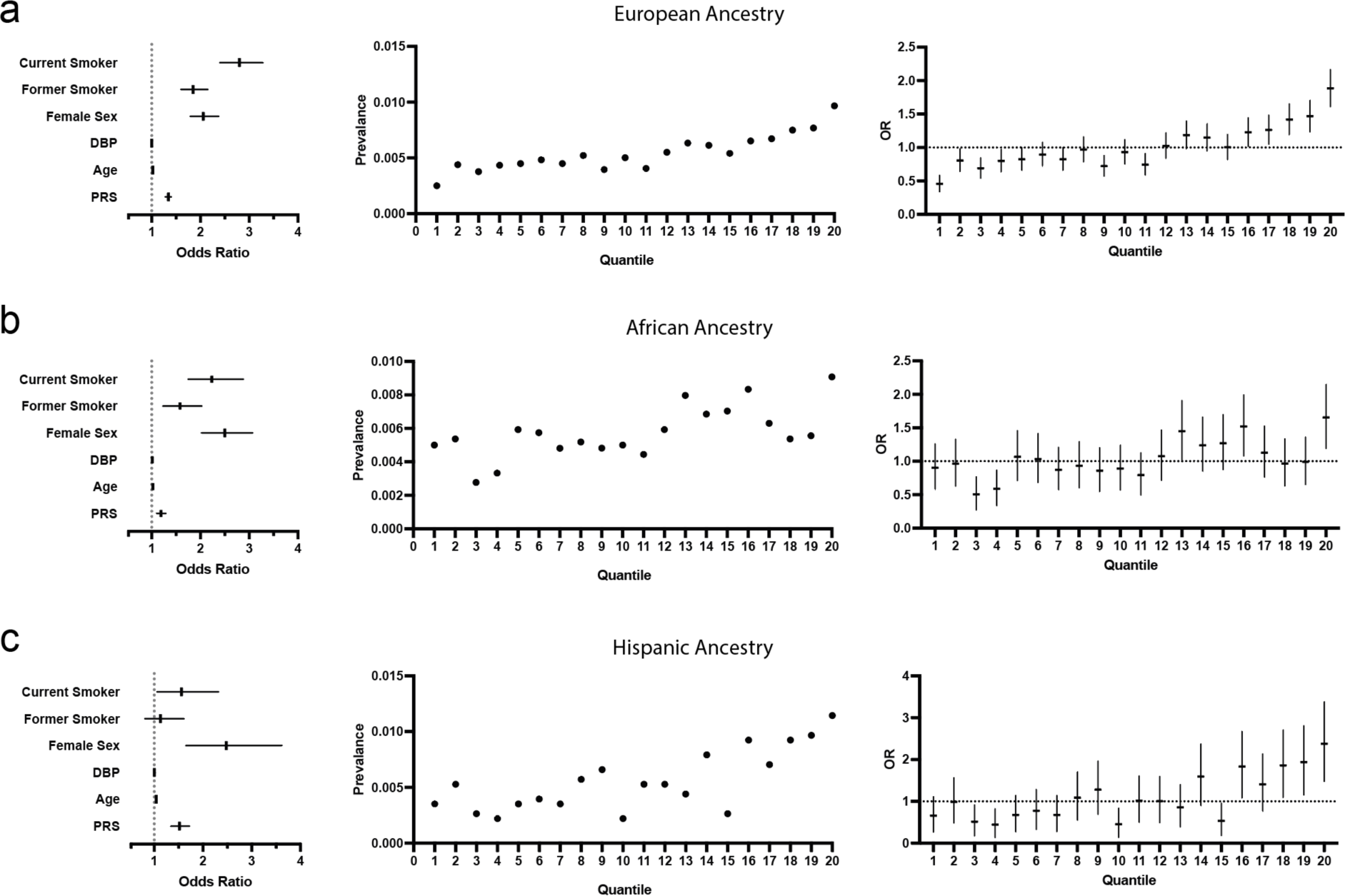
Generation of a Polygenic Risk Score to Predict Intracranial Aneurysm Diagnosis. Evaluation of a PRS in a) European b) African and c) Hispanic ancestries. Risk of IA diagnosis increases with higher polygenic risk scores (standard deviation unit) controlling for demographic parameters (left). Prevalence (middle) and odds ratio (right) of IA increases with PRS ventile.

